# Sharp/Needlestick Injuries Among Clinical Students at A Tertiary Hospital in Eastern Uganda

**DOI:** 10.1101/2023.02.01.23285330

**Authors:** Ndyamuhakyi Elisa, Lydia Ssenyonga, Jacob Stanley Iramiot, Doreck Nuwasiima, Rebecca Nekaka

## Abstract

**Background:** Clinical students, like health workers, are at risk of sharp/needle stick injuries and potential percutaneous exposure to blood and body fluids. They acquire infections like Human Immunodeficiency Virus (HIV) and Hepatitis B Virus (HBV) through injuries. This study determined the prevalence and factors associated with sharp injuries among clinical students at Mbale Regional Referral Hospital.

**Methods:** Across sectional study was carried out at Mbale Regional Referral Hospital, a teaching hospital located along Pallisa road, Mbale City, Eastern Uganda. Ethical approval was obtained, Printed and soft copy questionnaires eliciting demographics, injury occurrence and associated factors were randomly and conveniently distributed respectively to 322 clinical students. Data was entered in Microsoft excel, cleaned and exported to STATA version 14 for analysis.

**Results:** One hundred sixty (55.2%) clinical students had sustained a sharp injury in their clinical practice with a self-reported prevalence of 46.6% in the last year. The majority of the students 93(68.9%), sustained multiple sharp injuries. The common cause and site of injury were solid needles 72(45%) and finger (83.1%). Most students, 197(67.9%) reported ward procedures not being supervised and 124(42%) students worked on 15 and above patients daily. Final year clinical students were more likely to sustain sharp injuries than semi-finalists (P=0.000, OR 3.195% CI 1.7-5.5). Students who worked on ≥15 patients were more likely to sustain a sharp injury than those who attended to < 15 patients daily (P=0.000, OR 6.3 95% CI 3.7-10.8%). Students’ knowledge about sharp/infection control was limited.

**Conclusion:** This study showed a high prevalence of needle stick injuries among clinical students. The associated factors were; the year of study, having not learned about infection control, the number of patients attended to daily. Students should attend to a manageable number of patients, carry out procedures not rushing while supervised. It is important to create awareness and train students on infection control before and during their deployment in clinical areas as their health and the future of the health sector depend on them.

## Background

Globally, 3 million sharp injuries are estimated to occur among health workers in a medical setting every year (1). Health workers are often at risk of sharp and needle stick injuries and potential percutaneous exposure to blood and other body fluids (2). These injuries occur due to unsafe handling of sharp and needles during surgical operations, suturing, intravenous cannula insertion, intramuscular injections, blood sample collection, and disposal of the sharp and needles (3).

Early and recent studies show that sharp and needle stick injuries may lead to the transmission of serious chronic infections like Human Immunodeficiency Virus (HIV), Hepatitis B Virus (HBV), and Hepatitis C virus (HCV) (4) (5). The World health organization (WHO) estimated that over 40% of HIV and HCV cases in health workers resulted from sharp and needlestick injuries in 2002 (6). Over 90% of the infections resulting from sharp and needle stick injuries occur in developing countries due to resource-limited working conditions, lack of safety-engineered equipment, high prevalence of infectious diseases, and inadequate staffing of the health facilities (7).

Due to inadequate staffing, clinical students who desire to learn new procedures are actively involved in medical procedures (8). In their determination to learn new procedures, clinical students are likely to be at a higher risk for sharp and needle stick injuries (9). Studies done in Nigeria among medical students and Haramaya University in Eastern Ethiopia among midwifery and nursing students reported high prevalence of sharp and needlestick injuries of 48% and 62.8%, respectively (10)(11). It is also suggested that other factors like underdeveloped manual skills, limited clinical experience, and less knowledge on infection prevention and control may also lead to increased occurrence of sharp and needlestick injuries among clinical students(9)(10). A study done in Mexico among medical students and interns revealed a high prevalence of 58.2% sharp injuries with students who are not trained on infection control and prevention twice more likely to sustain needlestick injuries than the trained ones (12)(5).

In Uganda, the few studies that have been done to identify the prevalence of sharp and needle stick injuries among clinical students have also reported a high prevalence. A study done in Mbarara regional referral hospital among medical students and interns reported a prevalence of 55% (13). A different study done among students of Kuluva comprehensive school of nursing and midwifery in Arua district, northern Uganda indicated that 25.3% of the nursing students sustained a needlestick injury, with more than 50% of the cases from potentially infectious sources (14). Moreover, studies show that clinical students acquire infections through these injuries; for example, 11% of medical students at Makerere University, Kampala were infected with Hepatitis B virus through accidental sharp/needlestick injuries in 2005 (15). In the course of their study, clinical students are at high risk of sustaining sharp and needle stick injuries and potentially acquiring serious blood-borne infections such as HBV, HCV, and HIV. Also, students suffer psychological effects such as stigma from colleagues, anxiety, depression, and side effects of anti-retroviral drugs such as nausea, vomiting, and hallucinations. This strongly affects them academically, socially, financially, and in practice. In the last 10 years, more training institutions for nurses and doctors have opened up in Uganda. However, the prevalence of sharp and needle stick injuries among students in these institutions is not known. Busitema University Faculty of Health Sciences, Islamic University in Uganda, Mbale school of clinical officers, train doctors, nurses, anesthetists, and clinical officers at MRRH. The extent of sharp and needle stick injuries among the students training at this hospital is not known. Injuries among clinical students could even be higher. Thus, this study aimed at determining the prevalence and factors associated with sharp and needlestick injuries among clinical students at Mbale regional referral hospital.

## MATERIALS AND METHODS

### Study design

A cross-sectional study was carried out for a period of one month from September to October 2020.

### Study area

The study was carried out at Mbale Regional Referral Hospital, a teaching hospital located along Pallisa road, Mbale City, Eastern Uganda. Medical institutions training students at MRRH included Busitema University Faculty of Health Sciences, Islamic University in Uganda, Mbale college of health sciences, and Mbale School of nursing and midwifery. These institutions are located in Mbale city. Mbal city is located in the Eastern part of Uganda, lying between latitudes 1.08 and longitudes 34.175, covering an estimated area of 2,467km^2^.

### Study population

The study population was clinical students rotating at Mbale regional referral hospital. Clinical students are medical doctor, clinical officer, nursing and midwifery students who were in their practicum training/rotations at the hospital. These were from Busitema University, Islamic University of Uganda, Mbale College of health sciences, and Mbale School of nursing and midwifery..

### Sampling Technique

For hard copy questionnaires, simple random sampling was used where participants were chosen by assigning numbers from one to 30 till the sample size is reached whereas for soft copy questionnaires, convenient sampling was used where questionnaire link was sent to students whom we accessed their social media accounts.

### Sample selection Inclusion Criteria

Clinical students rotating at Mbale regional referral hospital and have consented to participate in the study.

### Exclusion criteria

Clinical students who were rotating at MRRH for their first time placement.

### Data Collection tool

The structured questionnaire both printed and soft copy was used to collect data. The Questionnaire was pretested in 10 intern students at MRRH. The online soft copy questionnaire linkwas sent to students through social media platforms like WhatsApp accounts, email addresses, Phone SMS inboxes, and Facebook messengers. Informed consent was obtained before questionnaire administration. When universities and schools re-opened for finalists on 28^th^ September 2020, a set of printed questionnaires were issued to students who were rotating onwards. These were students who had not participated in the online survey.

We used 18 items to measure respondents’ knowledge level regarding infections transmitted through, post-exposure management, and prevention of sharp/needle stick injuries by using true and false options. We selected 11 items to measure perception about sharp, infections transmitted, control, and prevention. We used five Likert scales (Strongly disagree, Disagree, Uncertain, Agree, and strongly agree) to measure the level of agreement on each selected item which was later condensed to Agree, Uncertain, and Disagree.

### Data management

Each questionnaire was given a code for identification. Completed questionnaires were downloaded kept on a password-protected computer and retrieved for processing by the researchers only. The hard copy-filled questionnaires were combined with the downloaded data set and cleaned. All data collected was double-checked for errors, cleaned, entered into Microsoft excel spreadsheet and exported to STATA version 14 for analysis. Coding was done by allocating numbers to particular responses from the participants to ease the analysis. All files were password protected and the password was only available to the researcher. Data were collected on demographics, occurrence, factors, knowledge, and perception of sharp/needlestick injuries.

### Data Analysis and Presentation

Data were analyzed using STATA version 14, summarized into percentages, frequencies, medians, means, and standard deviations where appropriate, and then presented in tables and figures like pie charts and histograms. Categorical data such as sex, course, year of study, procedure type, injuring previous object experience, sharp/needlestick injuries, work shift, was expressed in percentages and frequencies while continuous variables like age, number of patients (injections), and number of injuries were categorized. The prevalence, knowledge, and perception of sharp injuries were determined by univariate analysis, while associated factors were by bivariate and multivariate logistic regression.

### Ethical Considerations

Ethical approval was obtained from the Mbale Regional Referral Hospital Research Ethics Committee (MRRH-REC) under REC number MMRH-REC OUT 018/2020. Permission to collect data was sought from the administration of MRRH, Busitema University, Islamic University in Uganda, Mbale College of health sciences, and Mbale School of nursing and midwifery.

Written informed consent was obtained from all the participants and filled the questionnaires individually to ensure privacy, confidentiality, and anonymity. Participation was voluntary and participants could withdraw from the study at any time without penalty. There were no risks or monetary benefits to participating in the study. Clinical students who had got sharp/needlestick injuries before 72 hours had elapsed were advised to be assessed and given Post-exposure prophylaxis (PEP) as per the hospital and national infectious disease control guidelines.

## RESULTS

### Socio-demographic characteristics of respondents

Out of 322 students, 290 adult clinical students were enrolled in the study with a response rate of 90%. The majority of the participants (45.2%) were from Busitema University and 49.7% pursued a medicine course. Other participants were pursuing nursing, midwifery, or anesthesia courses. A majority (65.9%) of the participants were in the final year of study. The minimum and maximum age of participants was 19 and 43 years, respectively and the mean age 25.5 with a standard deviation of 4.7. The majorities (67%) of the participants were below 25years of age, about half were females and 48.6% were males. About three quarters (79%) of clinical students had been vaccinated against hepatitis B virus (Table 1).

**Table 1:** Socio-demographic characteristics of clinical students.

| Variables | Category | Freq. | Percentage |
| --- | --- | --- | --- |
| Age | 25 years and below | 194 | 67 |
|  | 26-30 years | 55 | 19 |
|  | Above 30 years | 41 | 14.1 |
| Gender | Male | 141 | 48.6 |
|  | Female | 149 | 51.4 |
| School | Busitema university | 131 | 45.2 |
|  | Islamic University in Uganda | 76 | 26.2 |
|  | Mbale college of health sciences | 47 | 16.21 |
|  | Mbale school of nursing/midwifery | 36 | 12.4 |
| Course | Medicine | 144 | 49.7 |
|  | Nursing | 139 | 47.9 |
|  | Anaesthesia | 7 | 2.4 |
| Year category | Finalist | 191 | 65.9 |
|  | Semi-finalist | 99 | 34.1 |
| Hepatitis B vaccination | Vaccinated | 61 | 21 |
|  | Not vaccinated | 229 | 79 |

### Prevalence of need stick injuries

Of the 290 students who participated in the study, 55.2% had ever sustained a sharp/needlestick injury, and 46.6% reported experiencing a sharp injury in the last year. Ninety-three (68.9%) students reported sustaining multiple sharp injuries. The most common injury site was the finger (83.1%) followed by the hand (16.9%). The most common cause of injury was solid sharp/needles 72(45%) followed by hollow needles. Injuries were sustained on all wards, with most occurring in emergency/causality ward 48(30%) followed by surgical ward 35(21.9%). Students were asked about various circumstances that could have contributed to needle stick injury and the majority 55(34.4%) reported rushing while doing the procedure as what to injury, whereas 40(25%) reported the patient being uncooperative, as shown in figure 1.

**Figure 1:**
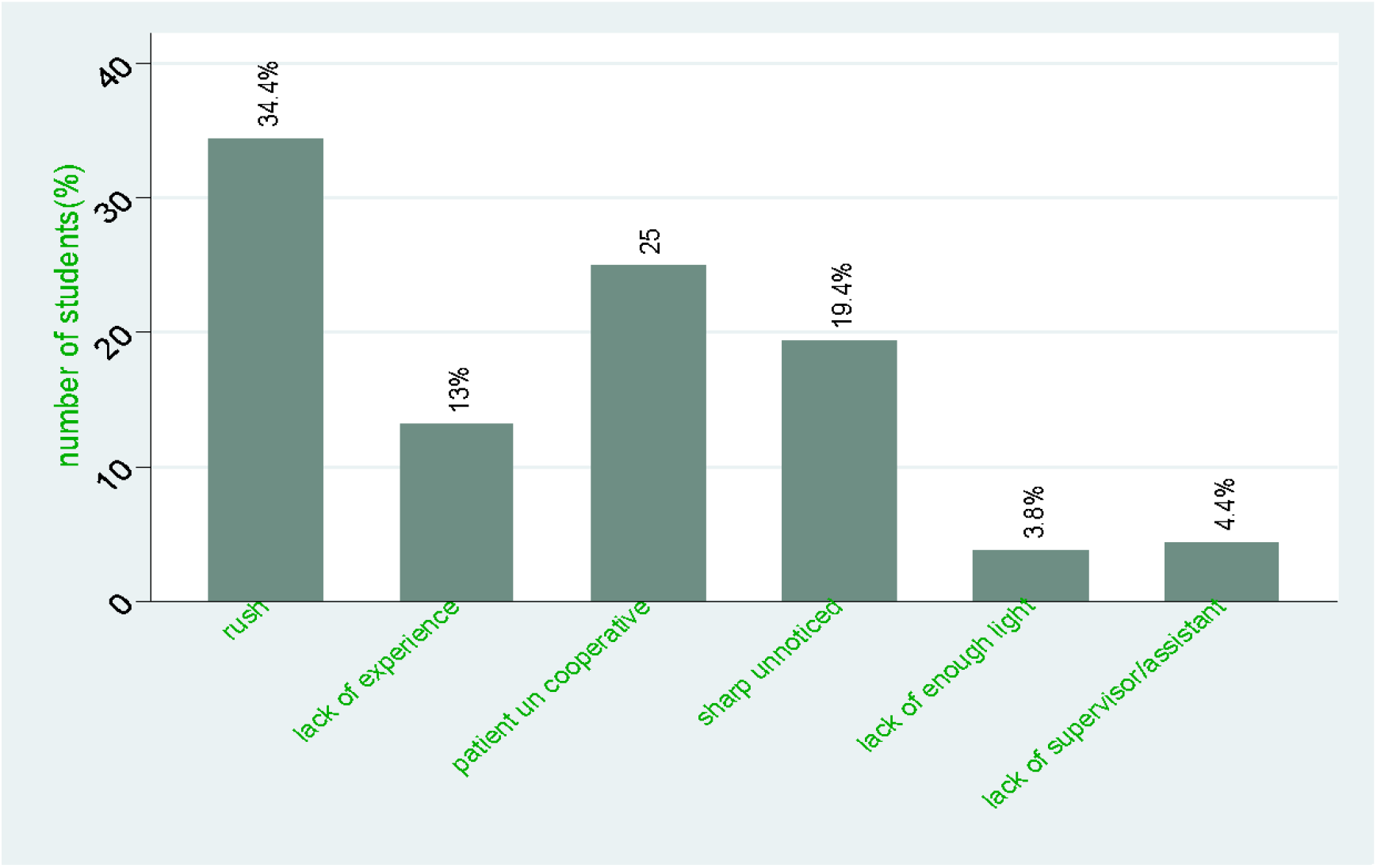
Reasons for sharp/needlestick injuries.

The majority of clinical students 45(28.1%) had a sharp injury while giving injections, followed by suturing 35(21.9%) as shown in figure 2.

**Figure 2:**
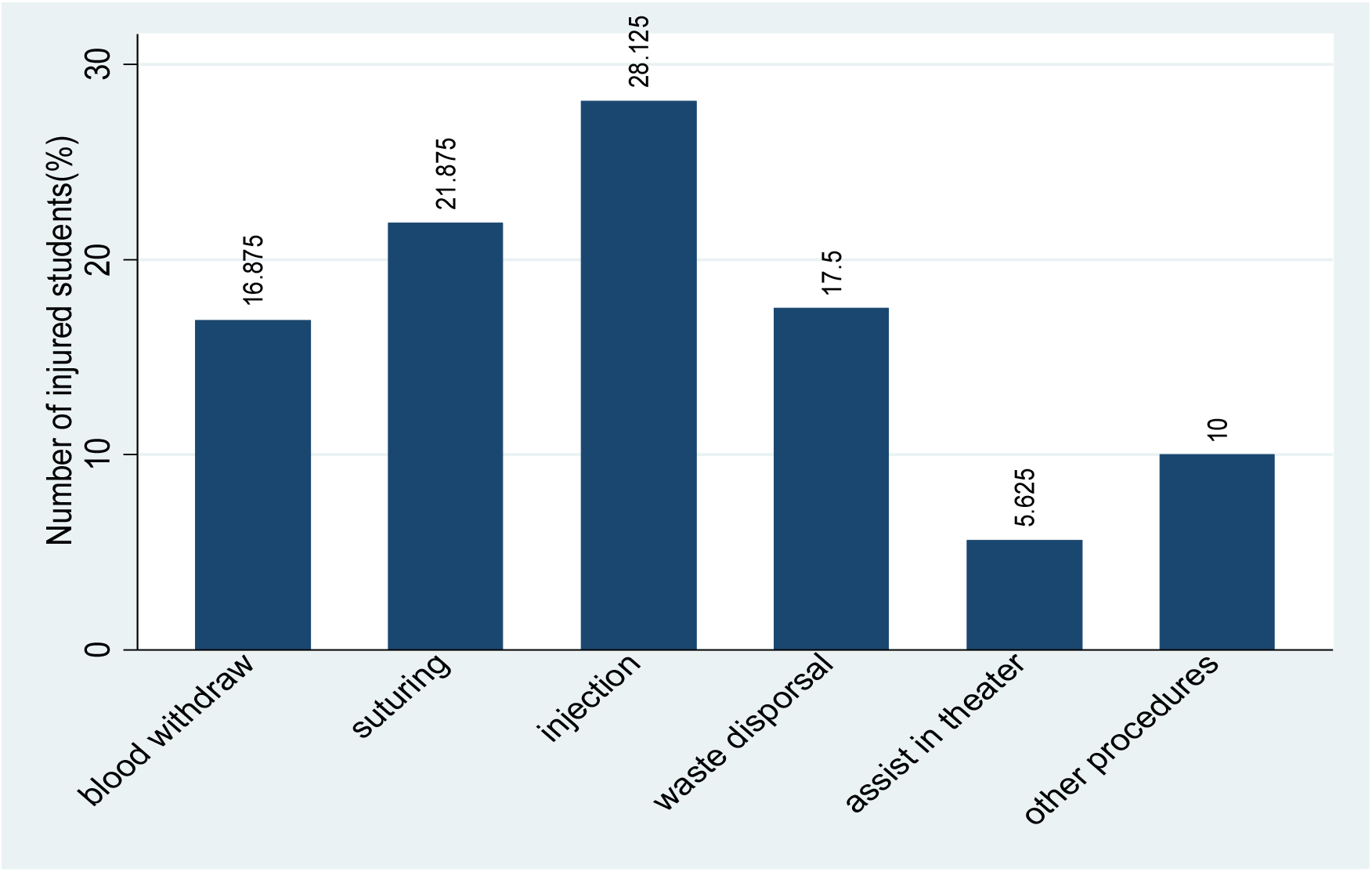
Procedures involved with sharp/needlestick injuries.

Most of the injured students (30%) were rotating in the emergency/casualty ward, then surgery (21.9%), and other wards. Students were asked about the actions they took after sustaining needle stick injury and only 54(47%) students reported having done all the three recommended measures as shown in Table 2 below.

**Table 2:**
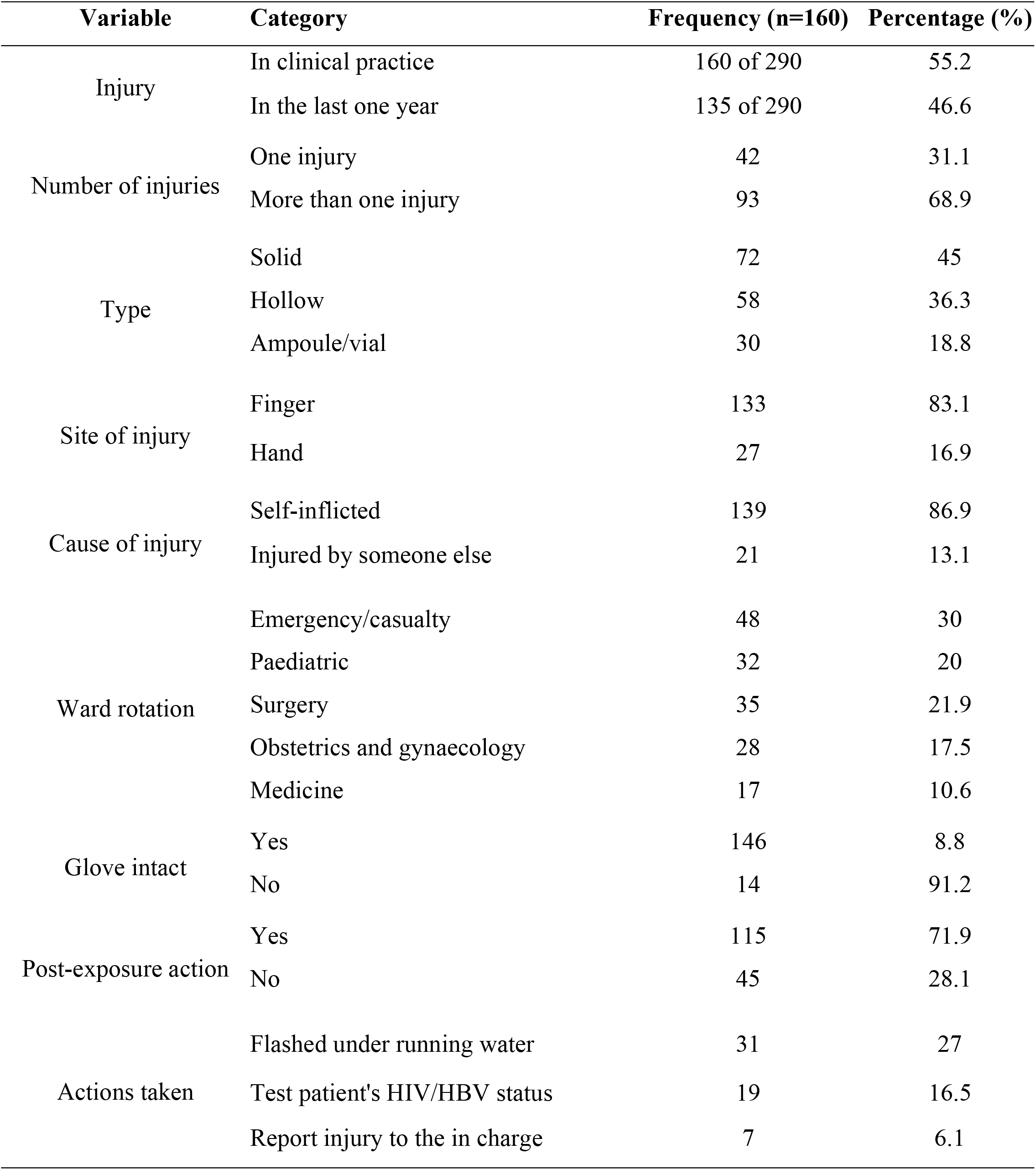
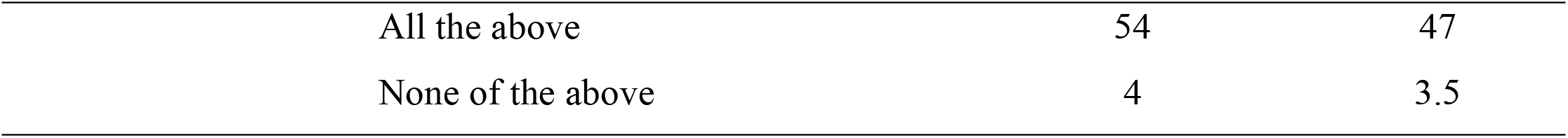
Information regarding injuries.

### Factors associated with sharp/needlestick injuries

In bivariate logistic regression analysis, school, course, year category, hepatitis B vaccination, ward procedure supervision, number of patients worked on daily, learning about sharp and infection control, were significantly associated with NSI with P<0.05 at 95% confidence interval. Only significant variables (P< 0.05) were entered for multivariate analysis. By adjusting potential confounders in multivariate logistic regression analysis, the only year of study, hepatitis B vaccination, the average number of patients attended to daily and having learned about sharp and infection control were significantly associated with sharp injuries. However, participant’s school, course, and ward procedure supervision were not significantly associated with sharp injuries in multivariate analysis.

Finalist clinical students were three times more likely to sustain a sharp injury than semi-finalist clinical students (P-value 0.000, OR 3.195% CI 1.7-5.5). Students who had not learned about sharp and infection control were two times more likely to sustain sharp/needlestick injury than those that had learned about sharp/infection control (P=0.012, OR 2, 95% CI 1.2-4). Students who attended to about 15 patients and above every day were six times more likely to sustain a sharp injury than those who attended to less than 15 patients (P=0.000, OR 6.3 95% CI 3.7-10.8%). Unvaccinated clinical students were 2.7 times more likely to sustain a sharp injury than those who were vaccinated (P=0.015, OR 2.7 95% CI 1.2-6) as shown in Table 3 below.

**Table 3:**
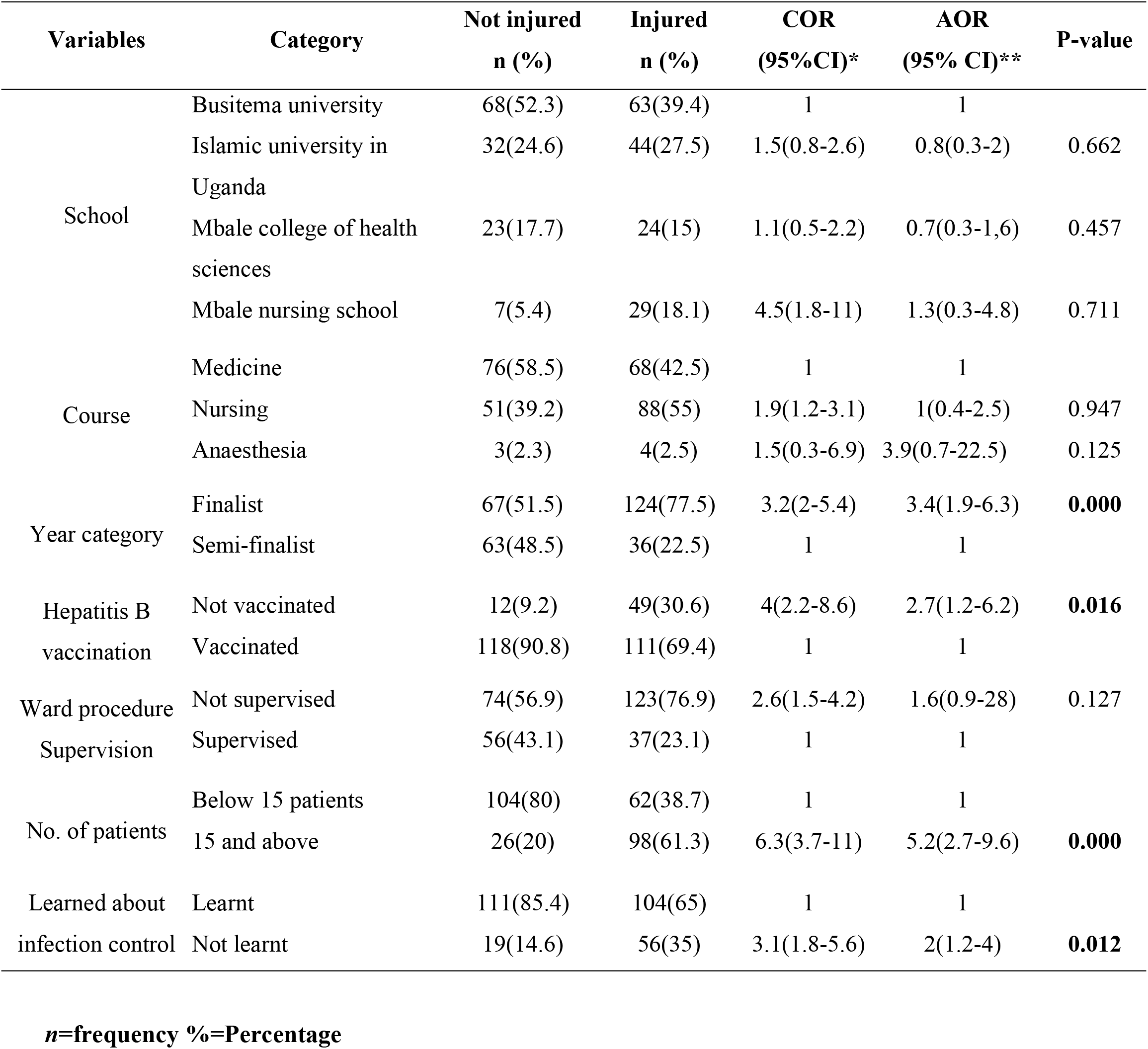
Factors associated with sharp/needlestick injuries.

**Table 4:**
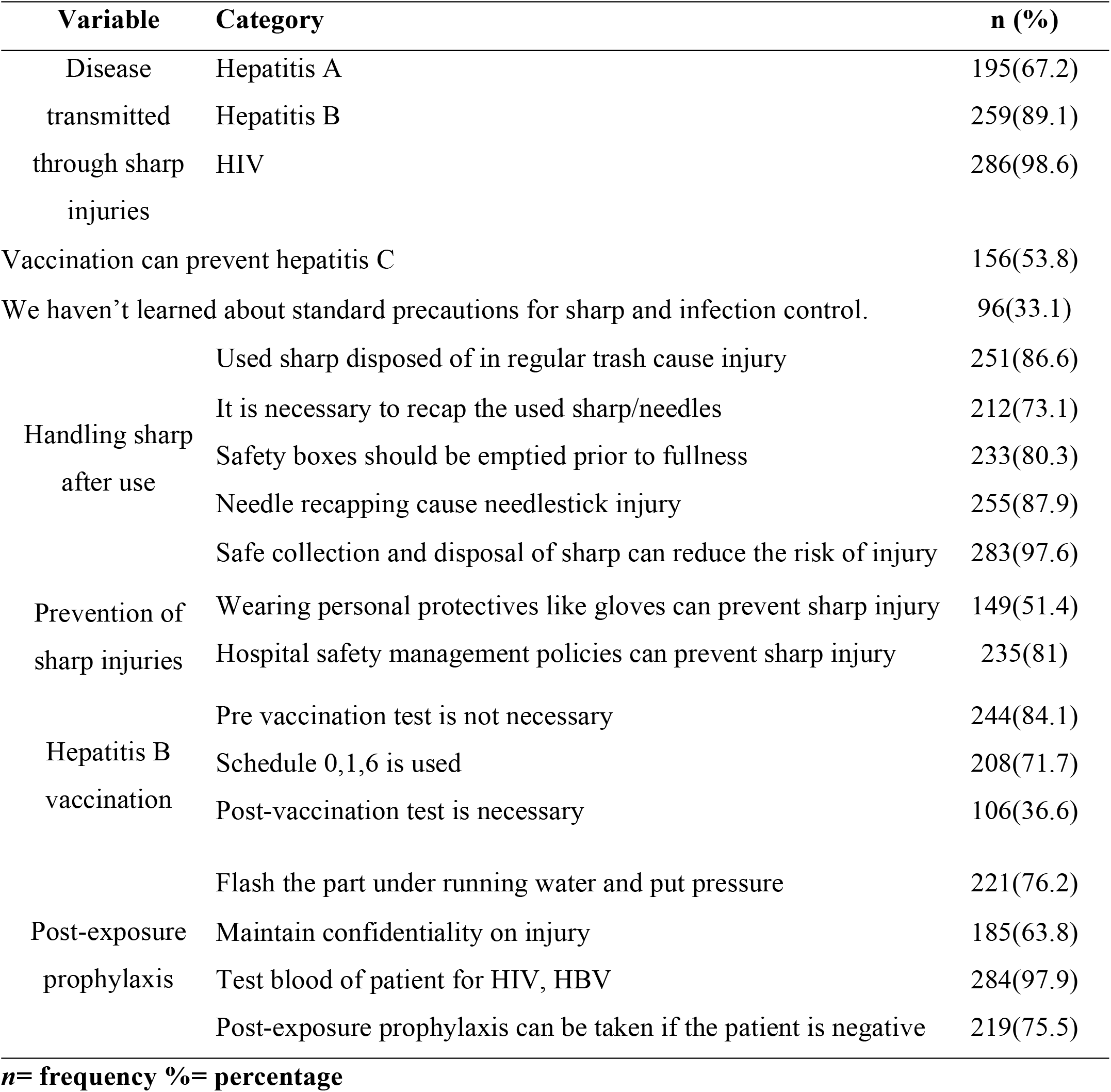
Knowledge concerning needle stick injuries (the correct responses)

### Knowledge concerning need stick injury

Regarding blood borne pathogens, only 67.2% knew that hepatitis A could not be transmitted through needle stick injury, 89.1% knew that hepatitis B could be transmitted through needle stick injury and 98.6% knew that HIV could be transmitted through need stick injury.

While handling sharp, 283(97.6%) reported that safe collection and disposal of sharp can reduce the risk of injury. Two hundred thirty three (80.3%) students agreed that safety boxes should be emptied before they are extremely full. Two hundred fifty-five,255(87.9%) students reported that needle recapping could cause sharp injury however only 212(73.1%) answered correctly if it was necessary to recap used sharp/needles.

In regards to preventing these injuries, 149 (51.4%) students agreed that wearing personal protective equipment like gloves for all patient care contacts is a useful strategy for reducing the risk of sharp injuries and, in turn, reducing transmission of infectious disease organisms. More so, 235 (81%) stated that safety management policies could prevent sharp injuries.

Regarding hepatitis B vaccination, the majority had knowledge regarding hepatitis B vaccination that is 44(84.1%) of the students correctly answered whether the pre-vaccination test was not necessary and 208(71.7%) knew that schedule 0, 1, 6 was used. However, only (36.6%) students believed that -post-vaccination test was necessary in relation to hepatitis B immunization. Only 53.8 % of the students knew that Hepatitis C infection could not be prevented by vaccination.

*On post-exposure prophylaxis measures, 221(76.2%) students reported flashing the part under running water and applying pressure to arrest bleeding as post-exposure action while 105(36.2%) agreed that after sustaining a sharp injury, they needed to report the injury to ward in charge. About 71(24.5%) clinical students agreed that post-exposure prophylaxis drugs could be taken even if the source patient was negative while 284(97.9%) students believed to test the blood of patient for HIV, HBV (Table 3). However, among the students who sustained a sharp injury, only 16.5% tested the patients’ HIV/HBV status, 27% flashed the injured part under running water, 6.1% reported the injury to the in-charge, whereas only 54% did all the recommended measures*.

### Perception of clinical students towards sharp/needlestick injury

Clinical students agree to the statement “Every health professional student has a chance to get sharp/needle stick injury” 267(92.1%). Two hundred fifty-seven (88.6%) also agree that “increased workload can lead to sharp/needle stick injury” though 238(82.1%) clinical students disagree that “If health care workers get infected with HIV infection, they should resign from their profession”.

Two hundred seventy-one, 271(93.4%) clinical students agreed that “The standard precautions to handle sharp objects must always be followed as improper handling can lead to getting the infection” and 237(81.7%) believed that “The infections transmitted from sharp/needle stick injuries are life-threatening.” Regarding this statement, “We haven’t learned about standard precaution for sharp/needle stick injuries”, only 25.9% agreed; however, 90.4% believed that “Confidence and skillfulness can prevent needle stick injury although there is a risk of infection.”

Most of the students 82.1% agree that “Unavailability of protective equipment can predispose a person to get sharp/needle stick injuries” and 231(79.7%) believed that “Handling sharp/needles without wearing gloves is better than wearing gloves”. However, 234(80.7) students disagreed that “Reporting after a sharp/needle stick injury is not useful.”

More than three-quarters 96.6% of students stated that “Every health professional student should be immunized against Hepatitis B infection.” Two hundred sixty-two (90.3%) students believed that “Health education for universal precaution on needle stick injuries to clinical students can reduce the prevalence of sharp/needle stick injuries among them” (Table 5).

**Table 5:** Perception responses about susceptibility to sharp injuries.

| **Perception** | **SA/A**<br>**n(%)** | **U**<br>**n(%)** | **D/SD**<br>**n(%)** |
| --- | --- | --- | --- |
| Every health professional student has a chance to get sharp/needle stick injury. | 267(92.1) | 7(2.4) | 16(5.5) |
| Increased workload can lead to sharp/needle stick injury. | 257(88.6) | 17(5.9) | 16(5.5) |
| If health care workers get infected with HIV infection, they should resign from their profession | 17(5.8) | 35(12.1) | 238(82.1) |
| The standard precautions to handle sharp objects must always be followed as improper handling can lead to getting the infection. | 271(93.4) | 6(2.1) | 13(4.5) |
| The infections transmitted from sharp/needle stick injuries are life-threatening. | 237(81.7) | 7(2.4) | 46(15.9) |
| Although there is a risk of infection, confidence and skilfulness can prevent needle stick injury. | 262(90.4) | 12(4.1) | 16(5.5) |
| Unavailability of protective equipment can predispose a person to getting sharp/needle stick injuries. | 238(82.1) | 15(5.2) | 37(12.7) |
| Handling sharp/needles without wearing gloves is better than wearing gloves. | 34(11.7) | 25(8.6) | 231(79.7) |
| Reporting after a sharp/needle stick injury is not much useful. | 51(17.6) | 5(1.7) | 234(80.7) |
| Every health professional student should be immunized with the Hepatitis B vaccine. | 280(96.6) | 3(1) | 7(2.4) |
| Health education for universal precaution on needle stick injuries to clinical students can reduce the prevalence of sharp/needle stick injuries among them. | 262(90.3) | 7(2.4) | 21(7.3) |
| **SA (Strongly agree), A (Agree), U(uncertain), A(Agree), SD (Strongly Disagree** |  |  |  |

## DISCUSSION

Health care worker trainees in pursuing their profession, especially nurses, medical and anesthesia students, face stressful clinical events placing them at an increased risk of needlestick injury.

### Prevalence of sharp/needlestick injuries

Our study found that 160 (55.2 %) clinical students had sustained sharp/needlestick injuries in their clinical practice. These findings agree with studies carried out among Nursing and Midwifery Students at Haramaya and Jigjiga University in eastern Ethiopia and among nursing students in Najing China that reported a high prevalence of sharp/needle sticks injury of 64.8% and 60.3% respectively (10)(16). However, our study findings were contrary to the study conducted in Bangalore India where only 25% of students experienced a sharp/needlestick injury (17). Our study finding is higher than that reported from a study conducted among nursing students in Uganda, where a low prevalence of 25% was reported (14). The difference could be a result of a big sample size used in our study compared to the one used in Hulme P, 2009. This high prevalence of sharp injuries injury could be attributed to limited experience on procedures carried out a factor Identified to be associated with sharp injuries(12). Also, possibly because of inadequate ward procedure supervision and many patients attended per day.

Findings from our study reveal a high one-year self-reported prevalence of sharp/needlestick injury of 135(46.6.4%). This finding is compared with a report from India where 36.5% of students had a sharp injury in the last one year (17). However, our study finding is lower than a cross-sectional study carried out in Ethiopia where students experienced a sharp/needlestick injury in the last one year, 62.8% (10). The high prevalence from our study could be due to poor techniques and skills about the procedure performed and non-adherence to standard precautions a factor which has been reported to be associated with the occurrence of needle stick injuries (14). Other reasons for this could be work overload, recapping of needles, and lack of personal protective equipment. However, a lower one-year self-prevalence of 11% and 13.9% have been reported in studies conducted in the UK and Australia, respectively (18)(19). This low one-year self-reported prevalence could be attributed to the presence of adequate personal protective equipment like gloves, Safe needle handling practices, safe and proper waste disposal in UK and Australia compared to developing countries like Uganda.

### Factors associated with sharp/need stick injuries

According to this study, 75 (25.9%) clinical students had not learned about infection control, among which 74.7% sustained a sharp/needlestick injury. Students who had not learned about sharp and infection control were two times more likely to sustain sharp/needlestick injury than those that had learned (P=0.012, OR 2, 95% CI 1.2-4). Similar findings were reported in a study conducted in Ethiopia where nursing students who had learned about infection prevention were 0.44 times less likely to sustain a sharp injury than those that did not learn (10). Studies done in Mexico and China also report that students not trained on sharp and infection control where twice likely to sustain a sharp injury compared to those that were trained (P=0.04 OR 3.21, 95% CI 1.44 -7.17) and (OR 2.1, 95% CI 1.4-3.2) respectively (12)(16). Students, while in their clinical training are in a daily encounter with sharp and infections thus students need to be trained on handling sharp and infection control before and during their ward training.

Another important factor was the year of study. Most of the sharp injuries occurred in finalist clinical students 124(77.5%) compared to semi-finalist 36(25.2%). Finalist clinical students were 3 times more likely to sustain sharp/needlestick injuries than semi-finalists (P-value 0.000 OR 3.1 95% CI 1.7-5.5). Similar findings were reported in a study done among dental undergraduate students at Ajman University of Science and Technology in the United Arab Emirates, where 66% of injuries occurred among finalist students(20). Moreover, Derek R. Smith, 2005, reported that needlestick injuries were most likely to happen in third (final) year nursing students compared to second and first-year counterparts. This might be because practical clinical experience culminates in the final year of study and these, students are gradually exposed to more needles and sharp activities as their competency improves and their clinical skills develop. This study thus suggests that there is a need for continued emphasis on sharp and infection control during students’ hospital practicum. However, being in the final year showed no significant association with needlestick injuries association in a study done in Ethiopia P=0.833,95% CI 0.4-3.6 (10). This disparity could be due to different training conditions students are subject to in these different countries.

In this study, the prevalence of sharp/needlestick injuries among students who worked on 15 patients and above was six times greater than those who worked on less than 15 patients (P=0.000, OR 6.3 95% CI 3.7-10.8%). However, this is contrary to a study conducted among Ugandan health workers where a high rate of sharp workers was found in health workers attending to less than 35 patients per day compared to those attending to more than 35 patients (21). This is possible because students are not experienced in handling many patients daily compared to already qualified health workers. More so 35 is still a high number of patients compared to 15 patients and above. However, it is possible in our study that those who worked on above 15 patients were at high risk of sharp injuries due to the invasive procedures they carried out. A study done among Chinese nursing students revealed that the night shift was a risk factor for sharp injuries. Nursing students who reported three or more night shifts per week were nearly six times more likely to experience sharp/needlestick injuries than those who did not work night shifts. This may be explained by lack of sleep resulting in inattention and less ability to concentrate when these students are providing treatments (22). Poor quality sleep has been cited as a contributing factor of sharp injuries among nursing students(16). Work overload resulting from working on many patients per day can also lead to the inability to concentrate and be attentive during procedures, thus likely to sustain a sharp injury. Our study, therefore, suggests that reduced workload may reduce the likelihood of sharp injuries.

Seventy-nine percent of students were completely vaccinated against Hepatitis B infection. These findings are high compared to a similar study conducted in Pakistan where almost two-thirds of health workers were not completely vaccinated against hepatitis B infection (23). This may be reflected due to poor accessibility, affordability, and prioritization between these countries’ health systems. Although the majority of students were vaccinated in this study, students who were not/partially vaccinated against hepatitis B were almost 3 times at greater risk of experiencing a sharp injury than those who were not vaccinated (P=0.016, OR 4 95%CI 1.2-6.2). Most unvaccinated students don’t know the dangers of sustaining needlestick injury and diseases transmitted and thus are careless while handling sharp and, in turn, experience a needle prick injury. Thus this study suggests that students should be immunized against Hepatitis B infection before enrolled for their clinical training.

### Knowledge on sharp and needlestick injuries

The majority of clinical students could correctly identify diseases transmitted through sharp/needlestick injuries 259(89.1%) and 286(98.6%) responded correctly that Hepatitis B and HIV could be spread via sharp injuries, respectively. However, 67.2% of students could not recognize that Hepatitis A is not transmitted through sharp injuries. This was also seen in a study done among health workers in Nigeria where 86.2% of students were knowledgeable on infections transmitted through occupational exposures(22). This could be explained that students had not learnt or been trained on sharp/infection control which is a factor shown to be associated with sharp injuries.

While handling sharp, 87.9% of students of this study were aware that needle recapping can cause sharp/needlestick injuries, and 73.1% of participants knew that it was not necessary to recap sharp/needles. Recapping needles prior to disposal has been shown to increase the risk of experiencing a needle stick injury (16). Similarly, 81.1 % and 70 % of students knew that used needles should never be recapped is contrary to the standard precautionary guidelines, which predispose students to sharp injuries(22).

Regarding the prevention of sharp injuries, 51.4% agreed that wearing gloves can prevent sharp injury. This was also shown in a study done in Nigeria where (98.6 %) health workers reported that wearing gloves while handling sharp could reduce the risk of sharp injury(22). This study found that 81% of students knew that hospital safety management policies could prevent sharp injury. However, this was different in a study done mong medical students where only 42% of students knew and were familiar with hospital management policies (19).

Our study revealed that 84.1% of students knew that pre-Hepatitis B vaccination test was necessary and 71.7% also knew that schedule 0, 1, 6 was used. However, 65.4% of students did not know that post-vaccination test was necessary. This could be due to the fact that the post-vaccination test is rarely done in our health systems due to financial constraints, given it is essential thus, students have not seen it being done.

Regarding post-exposure actions taken, most of the students had knowledge on what steps to take after sustaining a sharp injury except that more than a half (36.2%) of clinical students believed that they should maintain confidentiality (don’t report) the sharp injury. This was also seen in a study carried out among the nursing students in Nanjing, China where 86.9% of the injured students did not report the injury (24). This is contrary to the infectious disease control guidelines that recommend that we report the injury immediately to a fellow staff or the ward in charge, be assessed for infection risk and managed as per WHO guidelines (6). In this study, 28% of the injured students did not take any post exposure actions. Among those that took post exposure actions, only 47% followed all the recommended guidelines. This study thus suggests that more emphasis on poste exposure measures is needed.

### Perception towards sharp/infection control

In our study, the majority of students agreed that every student has chance to get injury which was similar in a study done in Abdulaziz University for Health Sciences, Saudi Arabia where 65.1% students disagreed that sharp injuries are the least encountered in clinical practice (25). Different studies have shown high prevalences and incidences of sharp injuries among health workers and student trainees (10)(26).

Findings of this study show that most students had a positive perception towards the use of personal protective equipment, for example, 82.1% of students agreed that unavailability of personal protective equipment can predispose a person to getting a sharp injury. About 79.7% of the students disagree that handling sharp without donning gloves is better than wearing gloves. The use of personal protective equipment has been identified as a factor in reducing risk of a sharp injury (16). However, in a study among health workers in Uganda, revealed that the perceptions and attitude towards needlestick injuries did not significantly influence the occurrence of needlestick injuries (21).

Regarding prevention of sharp, 280(96.6%) agreed that every clinical student should be immunized against Hepatitis B disease as a strategy to prevent infection transmission. Among 290 students in this study, 79 students had been completely immunized against hepatitis B disease (table 3). More so 90.3% of students agreed that health education for universal precaution on needle stick injuries to clinical students can reduce the risk of sharp/needle stick injuries. All these are strategies students believe can reduce the likelihood of a sharp injury. In a study done among medical trainees, 188(54%) felt insufficient safety training regarding infection. Awareness, learning and training about sharp and infection control have reduced risk for needlestick injuries P=0.002 (12). However, there was no significant association between this perception and sharp injuries (27). Our study suggests that there is a need to sensitize students on universal precautions, which might reduce the likelihood of sharp/needlestick injuries.

### Study limitations

This study was carried out one teaching hospital which limits generalizability to all clinical students in the whole country Uganda.

The results could have been affected by recall bias as respondents were required to recall past experience. More so, clinical students could have reported only injuries they thought were potentially infectious.

The study was initially affected by the outbreak of Corona Virus Disease (COVID -19) where it was challenging to access study participants since all training institutions were instructed to send students home, but we resorted to online data collection until schools were opened for finalist medical students.

Nevertheless, this study provides useful information about sharp/needlestick injuries and infection prevention and control.

## Conclusion

This study showed a high prevalence of needle stick injuries among clinical students. The associated factors were; the year of study, having not learned about infection control, the number of patients attended to daily. Students should attend to a manageable number of patients, carry out procedures under supervsion. It is important to create awareness and train students on infection control before and during their deployment in clinical areas as their health and the future of the health sector depend on them.

## Recommendations

Carry out continuous sensitization/training of students on infection control before and during students’ practical training deployment.

To ensure students work on a manageable number of patients to avoid patient over load to students which leads to rushing that predisposes them to injuries.

Conduct regular supervision of students while carrying out ward procedures and use personal protective equipment, such as gloves, gowns, while carrying out procedures.

Ensure students are immunized against Hepatitis B before they are deployed on wards as it not only protects them but also sparkles them a need to strictly observe infection prevention measures.

Further research to ascertain the effects of sharp/needle stick injuries among clinical students during their training is recommended.

To achieve these measures, a joint collaboration and commitment from academic institutions and hospital authorities is needed to establish committees that aim at improving the safety of student trainees and promote compliance to safe work practices like use personal protective equipment.

## Data Availability

OSF: Sharp/Needlestick Injuries Among Clinical Students at A Tertiary Hospital in Eastern Uganda DOI 10.17605/OSF.IO/WFNMV

https://doi.org/10.17605/OSF.IO/WFNMV

## Acknowledgment

We acknowledge the administration of Busitema University nursing department, Islamic University in Uganda, Mbale College of health sciences and Mbale nursing and midwifery school and other stake holders who supported and provided us with information pertaining this research. We appreciate the immense contribution of the management and staff of Mbale Regional Referral Hospital who allowed us conduct this study on clinical students.

We acknowledge the clinical students from Busitema University, Mbale school of nursing, Mbale school of clinical officers and Islamic university in Uganda who participated in this study by filling the questionnaires.

